# Costs and models used in the economic analysis of Total Knee Replacement (TKR): A Systematic Review

**DOI:** 10.1101/2022.11.14.22282318

**Authors:** Naline Gandhi, Amatullah Sana Qadeer, Ananda Meher, Jennifer Rachel, Abhilash Patra, Jebamalar John, Aiswarya Anilkumar, Ambarish Dutta, Lipika Nanda, Sarit Kumar Rout

## Abstract

**Objectives:** The major objective of this review was to summarize the evidence on the core modelling specifications and procedures on the cost-effectiveness of TKR compared to non-surgical management. Another objective of this study was to synthesize evidence of TKR cost and compare it across countries using purchasing power parity (PPP).

**Methodology:** The electronic databases used were MEDLINE (PubMed), Cochrane Central Register of Controlled Trials (CENTRAL), HTAIn repository and Cost effectiveness Analysis (CEA) registry. Consolidated Health Economic Evaluation Reporting Standards (CHEERS) was used to assess the validity of the methods and transparency in reporting the results of the included studies. The cost of TKR surgery from high income and low-or middle-income countries were extracted and converted to single USD ($) using purchasing power parities (PPP) method.

**Result:** 29 studies were included in this review, out of which eight studies used Markov model, five studies used regression model, one study each reported Marginal structure model and discrete simulation model and decision tree analysis to assess cost-effectiveness of TKR. For PPP, 23 studies were included in the analysis of TKR cost. The average cost of TKR surgery was lowest in developing country like India ($3457) and highest in USA ($19,645).

**Conclusion:** The findings of this review showed that the Markov model was most widely used in the analysis of the cost effectiveness of TKR. Our review also concluded that the cost of TKR was higher in developed countries as compared to developing countries.

## Introduction

Osteoarthritis is a degenerative joint disease involving the cartilage and surrounding tissues and is the leading cause of disability worldwide among older adults (1). Globally, the prevalence of OA knee was estimated to be around 22.9% (2) whereas In India, it was estimated to be 28.7% (3). The direct cost of OA knee was US$5294 per person per year for those aged over 65 years and $5704 for patients less than 65 years. This was estimated to be twice of non-OA patients. The Indirect costs of OA knee was around US$4603 per person annually, mainly due to work-related losses and home-care costs. The Total knee replacement (TKR) is considered as one of the interventions to overcome the burden of OA knee. The number of TKRs being done to mitigate the burden of OA knee has also been increasing throughout the world. In the United States of America, in the year 2010, approximately 700,000 TKR surgeries were performed, and its demand is predicted to grow to 3.8 million per annum by the year 2030 (4).

The cost of TKR in developed countries - USA is around $17500 (in 2017) and in European countries - Denmark and UK are €13149 (in 2020) and £7313 (in 2013) respectively, however, in developing countries like India, it is ₹80,000 (in 2021) (5–8). Many developed countries have considered cost-effectiveness analysis as one of the methods for policy level decision-making. However, India, which has multiple health system constraints and limited government investment on health, is progressively preparing to include cost-effective analysis as a tool for decision-making at the policy level. There has been only one study conducted in India, which showed that TKR is cost-effective in the base case scenario with an Incremental Cost Effectiveness Ratio (ICER) of ₹9789 ($132.3) per QALY(9).

Given the emerging disease burden of osteoarthritis in both developed and developing countries and its associated interventions involving huge financial burden both to the individuals and government, it is critical to review the current literature regarding the cost effectiveness of such interventions and impact of the prevailing cost trends on the health systems. Moreover, given varieties of models-the decision tree, Markov model, and regression analysis (10) used in these types of studies, identifying the suitable model also assume significance. During our literature search, four systematic reviews were identified which compared the cost-effectiveness of TKR to non-surgical management in patients with OA knee. Two reviews compared the cost-effectiveness of TKR through ICER value irrespective of the models used (11,12). A recent review by Kamaraj et al (13) assessed the scope and quality of all current cost-effectiveness analysis (CEA) studies for TKR in order to identify trends, and recognise the areas for improvements. Further, another review by Lan et al (14) focused on the study designs and compared the Markov model with RCT and tree diagram in the analysis of cost-effectiveness of TKR.

However, none of the systematic reviews mentioned above suggested a suitable method for the analysis of cost-effectiveness of TKR. Therefore, we aim to perform a systematic review of the methods used in economic evaluations/cost-effectiveness of TKR compared to non-surgical management of OA knee. The major objective of this study is to summarize the evidence on the core modelling specifications and procedures on the cost-effectiveness of TKR compared to non-surgical management. Another objective of this study is to synthesize evidence of TKR cost and make different countries cost comparable using purchasing power parity (PPP), which has not been done before.

## Methodology

A systematic review protocol was registered in International Platform of Registered Systematic Review and Meta-Analysis Protocols (INPLASY) with registration no:(INPLASY202290044). Cochrane methodology were adopted and the Preferred Reporting Items for Systematic Review and Meta-analysis (PRISMA) guidelines were used for the purpose of reporting.

### Criteria for considering studies in the review

#### Types of Studies

In this review, all the reports of randomized control trials (RCT’s) and cohort studies were included. Studies like cross-sectional, observational and case-control that have the possibility to provide information on cost-effectiveness, cost benefit analysis, and cost utility analysis were also included in our initial search.

#### Types of Participants

Participants aged 40 years and above who have primary OA knee

#### Type of Intervention

Participants with OA knee who underwent surgical intervention that is total knee replacement (TKR).

#### Comparator

Participants with OA knee without surgical intervention (TKR) or underwent non-surgical management.

#### Types of Outcome Measures

Economic evaluation studies that report outcomes - Incremental Cost-effectiveness Ratio (ICER), cost-effectiveness, and improvement in QALY.

### Search Strategy for identification of the studies

The electronic databases included in the search were MEDLINE (PubMed), Cochrane Central Register of Controlled Trials (CENTRAL), HTAIn repository and CEA (Cost-effective analysis) registry. The search history was conducted from the earliest possible date till 4^th^ June 2021 and filters were not applied for time period. The computer-based search terms included the combination of keywords and Mesh terms like “Total Knee Arthroplasty” and “Cost-Effectiveness” ‘Knee osteoarthritis’, ‘Cost-utility analysis’, ‘Total Knee Replacement’ and ‘Economic Evaluation’ ‘Non-surgical Management” and comparators (NSAIDs, steroids, visco-supplementation, physiotherapy, exercise), and outcome (quality of life, EQ5D, ICER). Snowballing was also performed to identify other relevant articles. The search strategies for the electronic database are in the S1 File.

### Inclusion and Exclusion Criteria

1. Studies with English language were included
2. Studies with participants aged 40 years and above with degenerative OA knee were only included whereas traumatic OA knee-based studies were excluded.
3. Study design such as Randomized control trial or quasi-randomized control trial-based on TKR (+ Postsurgical management) study and/or Non-Surgical Management with Cost-effectiveness, cost benefit analysis, cost utility analysis along with prospective observational study on TKR (Post-surgical Management) and/ or non-surgical management were included.
4. Studies that report costing, Quality-Adjusted Life Years (QALYs), and economic models for calculating ICER of TKR, TKR (+post-surgical management), and TKR vs non-surgical management were included.
5. Studies considering other surgical procedures like Partial Knee Arthroplasty (PKA), Unicompartmental Knee Arthroplasty (UKA), and Kinespring Implant were excluded. Simultaneous bilateral TKR and revision TKR studies were also not included.
6. Studies were excluded if incomplete economic analyses like only cost comparison or only QALY comparison were reported. Non-economic evaluations were also excluded.
7. Studies such as systematic reviews, letters to the editors, commentaries, and protocols were excluded.

### Selection of Studies

All the identified studies from the database were imported to the RAYYAN software and duplicates were removed. During the “first pass”, three authors independently reviewed the study titles and abstracts before concluding whether the study should be “included,” “excluded,” or there is “uncertainty” about it. The consensus was reached in studies that had “uncertainty.” The remaining studies were excluded, while those that were “included” were considered for the next stage of selection.

Three authors retrieved the full texts of the selected abstracts from the first pass and screened them again. Similar screening methods were applied, but the authors agreed to include papers through an iterative consultation process for those with conflicting findings. Papers with unresolved conflicts were excluded after this process.

### Data Extraction and Management

The selected full text articles data were extracted for subsequent synthesis. A data extraction framework was developed for extracting data based on the criteria such as authors name, year, study location, type of model, perspective, direct and indirect cost of TKR and non-surgical management, ICER values, and QALYs. Data was extracted by two authors from the selected full-text papers, and it was verified by another two authors. The fifth reviewer was consulted for expert consensus in the event if there was any disagreement during the data synthesis.

### Conversion of Costing data on Purchasing Power Parities (PPP)

The cost of TKR differs across different metrics such as time and currency. Change in the prices due to inflation have affected the cost of TKR over the years. This study adjusted the cost of each country to 2021, using price index (inflation index) of different countries. Further, the cost from the high income and the LMIC countries were converted to single USD ($) using purchasing power parities (PPP) method (Cost in USD = Cost in any Currency / exchange rate in PPP). The PPP are the rates of currency conversion that equalize the power of different currencies(15)

### Quality Assessment of Studies

A 24-item tool was used to assess the validity of the methods and transparency in reporting the results of the study based on the Consolidated Health Economic Evaluation Reporting Standards (CHEERS) (16). The checklist was designed to qualitatively evaluate an economic study in terms of title and abstract, introduction, methods, results, discussion, disclosure of funding source, and conflict of interest. Two authors evaluated the studies based on these parameters from the checklist and further, it was verified by another two authors. Expert consensus was drawn from a fifth author wherever necessary. Each item of the study was assigned “Yes” if the information is completely reported, “Part” if partially reported and “No” if not reported. NA was reported if the item was “not applicable”. A score of 1 was assigned if the criteria was “Yes”, 0.5 if it was “Part” and 0 if the criteria “No”. Hence, the maximum and minimum score for a study were 24 and 0 respectively. Additionally, the percentage was determined under the presumption that each study received equal weighting, with “Not Applicable” item being left out. Studies with a score of at least 75% were defined as high quality, those between 60 and 75% as moderate quality, and those below 60% as low quality. The quality assessment did not influence the inclusion or the exclusion of studies. Studies meeting the inclusion criteria were included for the purpose of this review irrespective of the quality criteria.

### Data Analysis

Due to heterogeneity of the included studies and reported cost data, formal meta-analysis could not be performed. Thus, each study was analysed qualitatively and were primarily presented in tables and graphs.

As for PPP, out of the twenty-nine included studies, six studies were excluded from the analysis, of which three studies were excluded due to unavailability of cost data (17–19) and three studies due to being published before the year 2000(20–22), could not be included due to the data being older than 20 years. Studies conducted before the year 2000 were excluded from PPP analysis due to change in cost as the technology could be upgraded over the years. The remaining twenty-three studies(5–8,23–41) that were published after the year 2000 and presented direct cost of TKR were included in our analysis. The TKR cost of all studies was inflation adjusted to 2021 price in their respective country’s currency. The country wise inflation rate (in term of consumer price index) and the PPP value were extracted from world development indictors data base (published by World Bank)(42).

## Results

The search yielded total 801 articles (767 articles from the PubMed database, 23 articles from Cochrane central, 2 studies from CEA registry database and 9 from Google scholar). After removing duplicates (30 articles), 771 articles were eligible for title and abstract screening. After initial screening 77 articles were eligible for full text screening. Out of 77, full text was not available for 6 articles and 1 was not in English language, hence were excluded. Among the retrieved 70 full text articles, 41 articles did not meet the inclusion criteria. The reasons for exclusion being articles with different outcomes (12 articles), other study design (8 articles), intervention other than TKR (15 articles) and comparators other than non-surgical management like UKA, PKA (6 articles). Hence, 29 studies were included in this review for the descriptive synthesis. The process of selection of studies are mentioned in PRISMA flowchart (Fig 1).

**Fig 1:**
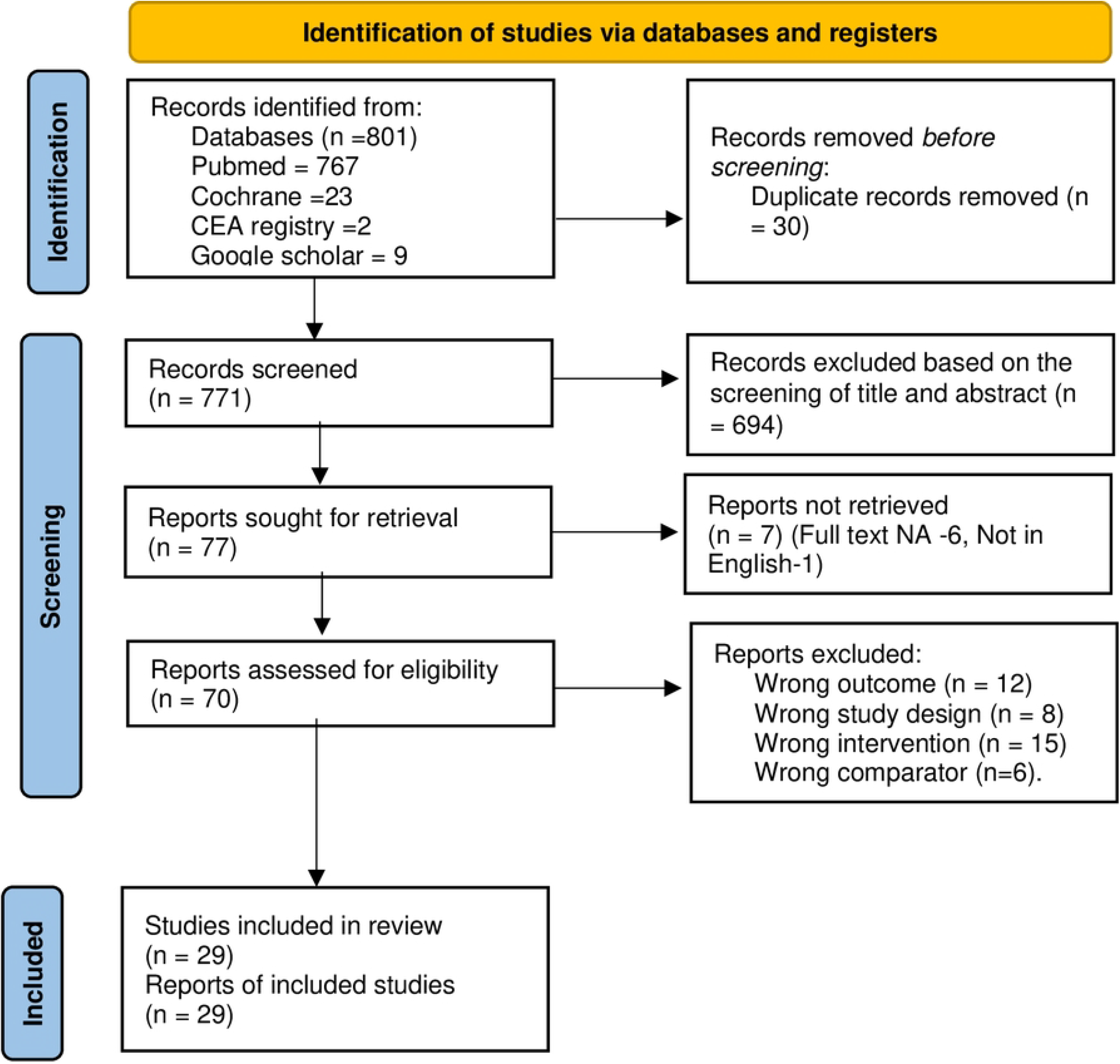
Studies selection process through PRISMA.

### Study Characteristics

#### Description of included studies

Twenty-nine studies (5–8,17–41) met the inclusion criteria and were published between 1993 and 2021. The included studies were from different parts of the world with eleven studies from United States (5,18,19,22–28,34), four studies from Canada (21,29–31), three studies from United Kingdom (8,35,36) and two each from Finland (17,20) and Australia (32,33). One study each was conducted in India (7), China (41), Germany (38), Spain (40), Romania (39), France (37) and Denmark (6) (See table 1).

**Table 1:**
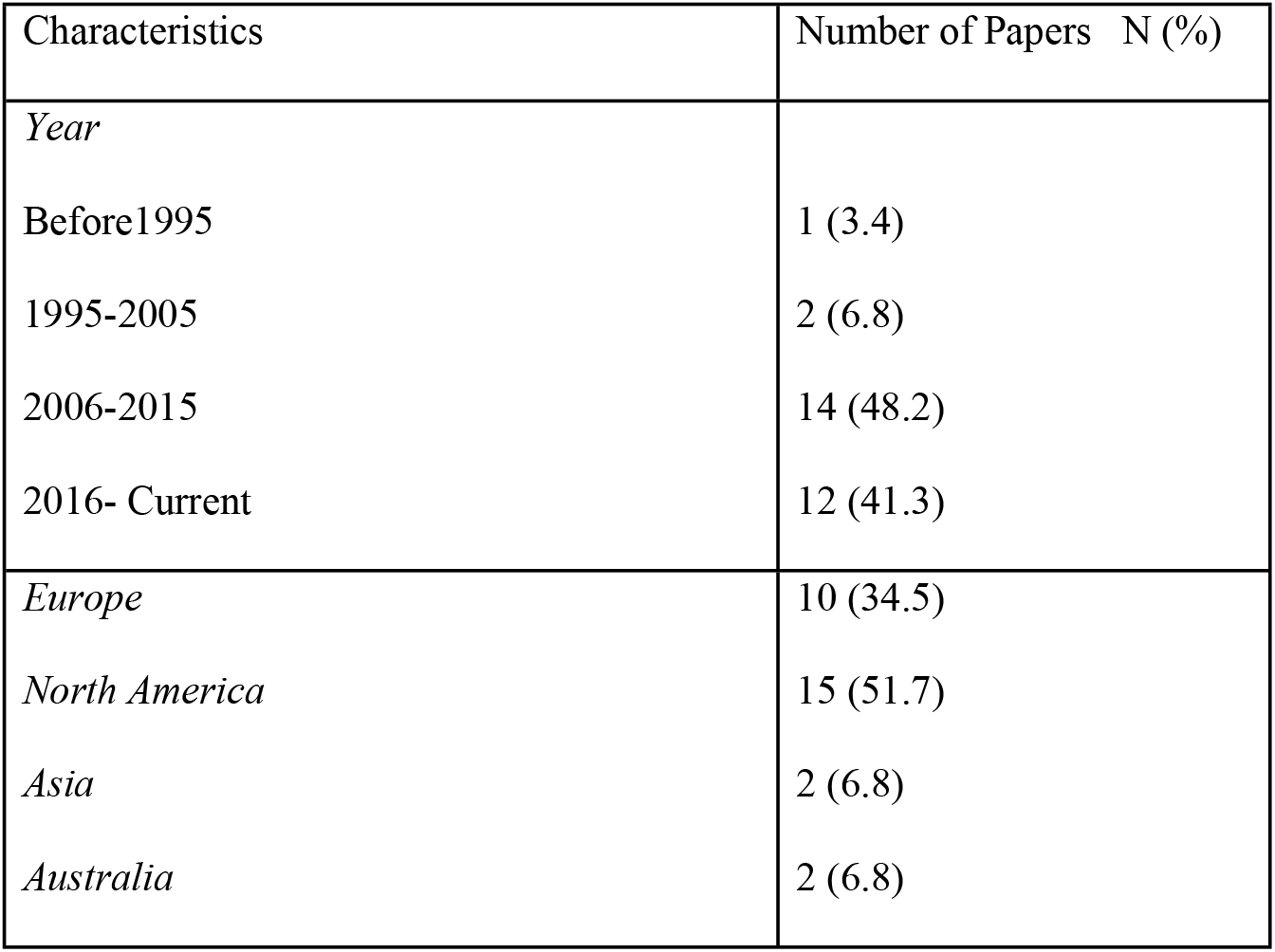
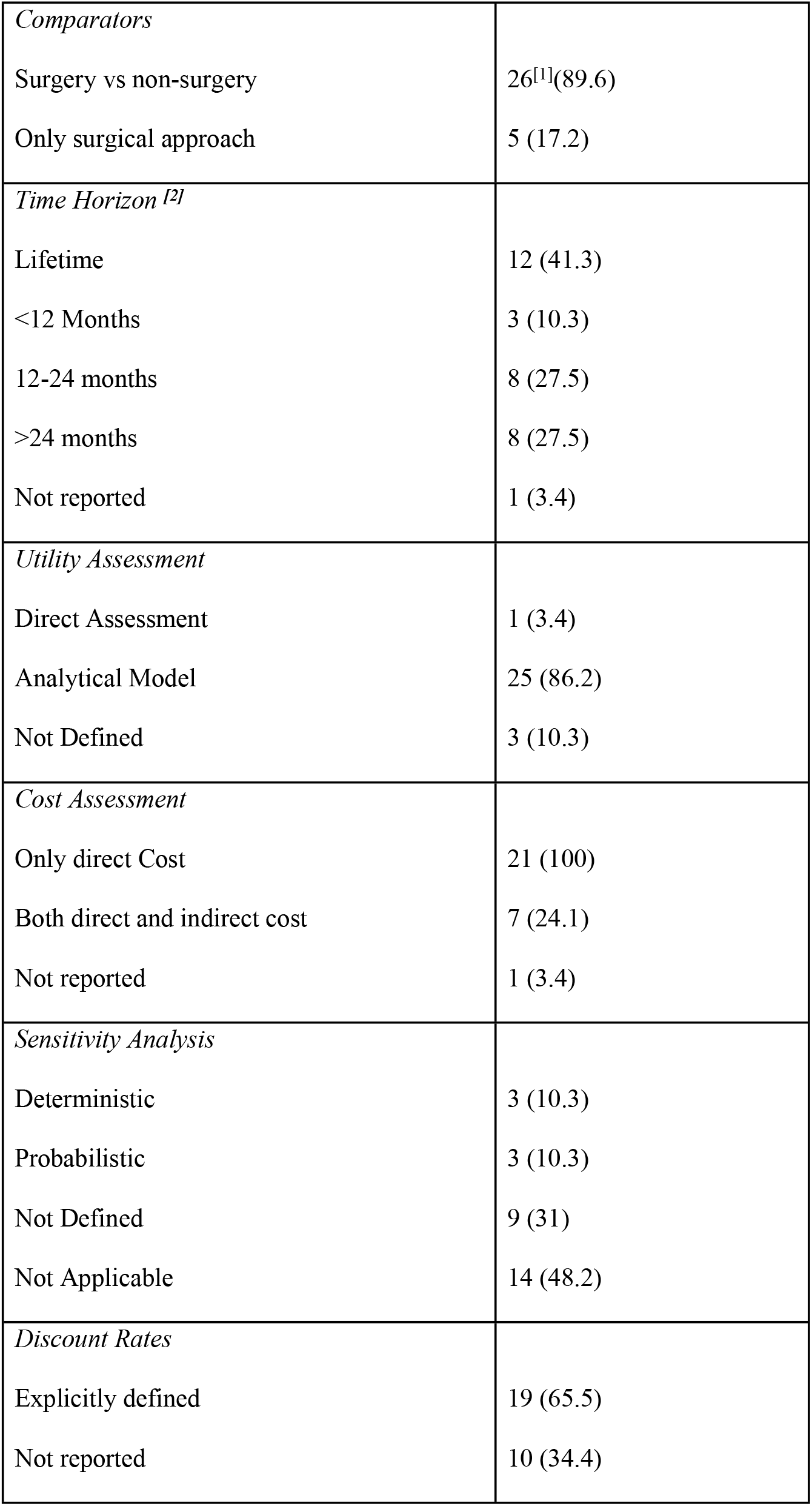
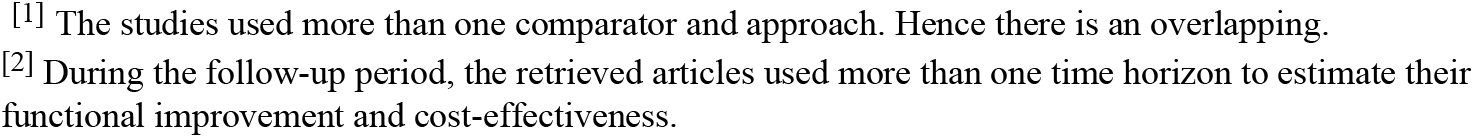
Characteristics of the selected papers.

| Characteristics | Number of Papers N (%) |
| --- | --- |
| *Year* |  |
| Before1995 | 1 (3.4) |
| 1995-2005 | 2 (6.8) |
| 2006-2015 | 14 (48.2) |
| 2016- Current | 12 (41.3) |
| *Europe* | 10 (34.5) |
| *North America* | 15 (51.7) |
| *Asia* | 2 (6.8) |
| *Australia* | 2 (6.8) |
| *Comparators* |  |
| Surgery vs non-surgery | 26<sup>[1]</sup>(89.6) |
| Only surgical approach | 5 (17.2) |
| *Time Horizon <sup>[2]</sup>* |  |
| Lifetime | 12 (41.3) |
| <12 Months | 3 (10.3) |
| 12-24 months | 8 (27.5) |
| >24 months | 8 (27.5) |
| Not reported | 1 (3.4) |
| *Utility Assessment* |  |
| Direct Assessment | 1 (3.4) |
| Analytical Model | 25 (86.2) |
| Not Defined | 3 (10.3) |
| *Cost Assessment* |  |
| Only direct Cost | 21 (100) |
| Both direct and indirect cost | 7 (24.1) |
| Not reported | 1 (3.4) |
| *Sensitivity Analysis* |  |
| Deterministic | 3 (10.3) |
| Probabilistic | 3 (10.3) |
| Not Defined | 9 (31) |
| Not Applicable | 14 (48.2) |
| *Discount Rates* |  |
| Explicitly defined | 19 (65.5) |
| Not reported | 10 (34.4) |

[1] The studies used more than one comparator and approach. Hence there is an overlapping. [2] During the follow-up period, the retrieved articles used more than one time horizon to estimate their functional improvement and cost-effectiveness.

Different types of methodologies were used to assess the cost-effectiveness of TKR in the included studies. Decision analytic model that is Markov model were used by eight studies (5,7,18,23,24,27,29,37) whereas five studies reported regression model (6,33,34,36,38). One study each reported a marginal structure model (28), discrete simulation model (32), and decision tree analysis (8) to assess the cost-effectiveness of TKR. Thirteen studies (17,19– 22,25,26,30,31,35,39–41) did not report any methodology to determine cost-effectiveness.

Ten Studies (7,8,19,23,24,27,28,32,38,39) considered time horizon to be life time whereas thirteen studies used ≥12 months of time horizon (5,6,17,18,21,22,29,30,33–36,41). Three studies (25,26,40) evaluated time horizon less than a year and one study(20) did not specify time horizon. Two studies (31,37) used more than one time horizon to estimate their functional improvement and cost effectiveness.

Nineteen studies reported discount rates, out of which, thirteen studies (6,9,19,23,24,27– 29,31,36,37,39,40) discounted cost and QALY at 2% to 6%. Two studies (5,18) discounted only cost at 3% and two studies (8,17) had discounted only QALY at 3.5% to 5%, while another study (32) discounted cost and Disability Adjusted Life Years (DALY) at 3%. Only one study (38) discounted benefit rather than cost at 3%. Two studies(26,35) did not apply discount rates and eight studies (20–22,25,30,33,34,41) did not report it.

Provider’s perspective was adopted in eleven studies (17,20,22,25,29–31,33,35,38,39) whereas five studies (23,24,26,27,34) reported the societal perspective. Few studies employed other type of perspective which are mentioned in Table-1.

### Model Structure and Outcome

Sixteen (5–7,7,8,18,23,24,27–29,32–34,36,37) out of twenty-nine studies used model to report outcomes like cost-effectiveness or cost-utility of TKR as mentioned in S2 file. Twelve studies did not provide any justification regarding the scope of model whereas Ferket et al (28) used marginal structure models for repeated measures and justified by stating that “outcome values can vary over time and can predict future treatment assignment along with other time-varying confounders”. Similarly, Higashi et al (32) justified the use of discrete event simulation (DES) model to be a better approach than Markov model since larger number of attributes and events are more likely to be managed by the DES model as opposed to the Markov model. In addition to this, it is possible to attach memories in previous states in a DES model, which is difficult to achieve in a Markov model. Dakin et al (36) used generalised linear model with gamma family distribution, as the costs and QALYs were highly skewed whereas Karmarkar et al (5) suggested that the objectives of the study could have been achieved through generalised linear model but the study used Markov model for its unique ability to forecast the societal cost of disparities as Markov model was widely used to determine cost-effectiveness of single treatment modality.

Clear and transparent modelling methodologies were presented in all the included sixteen studies. The source of data used to calculate transition probabilities were indicated in eight studies that used Markov model. Among these, seven included studies (5,7,23,24,27,29,37) incorporated transition probabilities from literature whereas only one study (18) used secondary data to calculate transition probabilities. All the included studies incorporated direct cost for the analysis whereas only seven studies (5,18,20,23,26,27,32) included indirect cost.

### Costing Data

Out of the 23 studies selected for Purchasing Power Parity analysis, eight studies were from USA (5,23–28,34), eight European countries -: from UK (8,35,36), Denmark (6), France (37), Germany (38), Romania (39) and Spain (40), three from Canada (29–31), two from Australia (32,33) and one from India (7) and one from China (41).

The cost of TKR surgery in India was found to be the cheapest at $3457 followed by China at $14,311. The average TKR cost in PPP ($) term was $16,123 in 2021. It is lowest in developing country like India ($3457) and highest in USA ($19,645). Region wise average cost of TKR surgery is presented in a bar graph as shown below in Fig 2.

**Fig 2:**
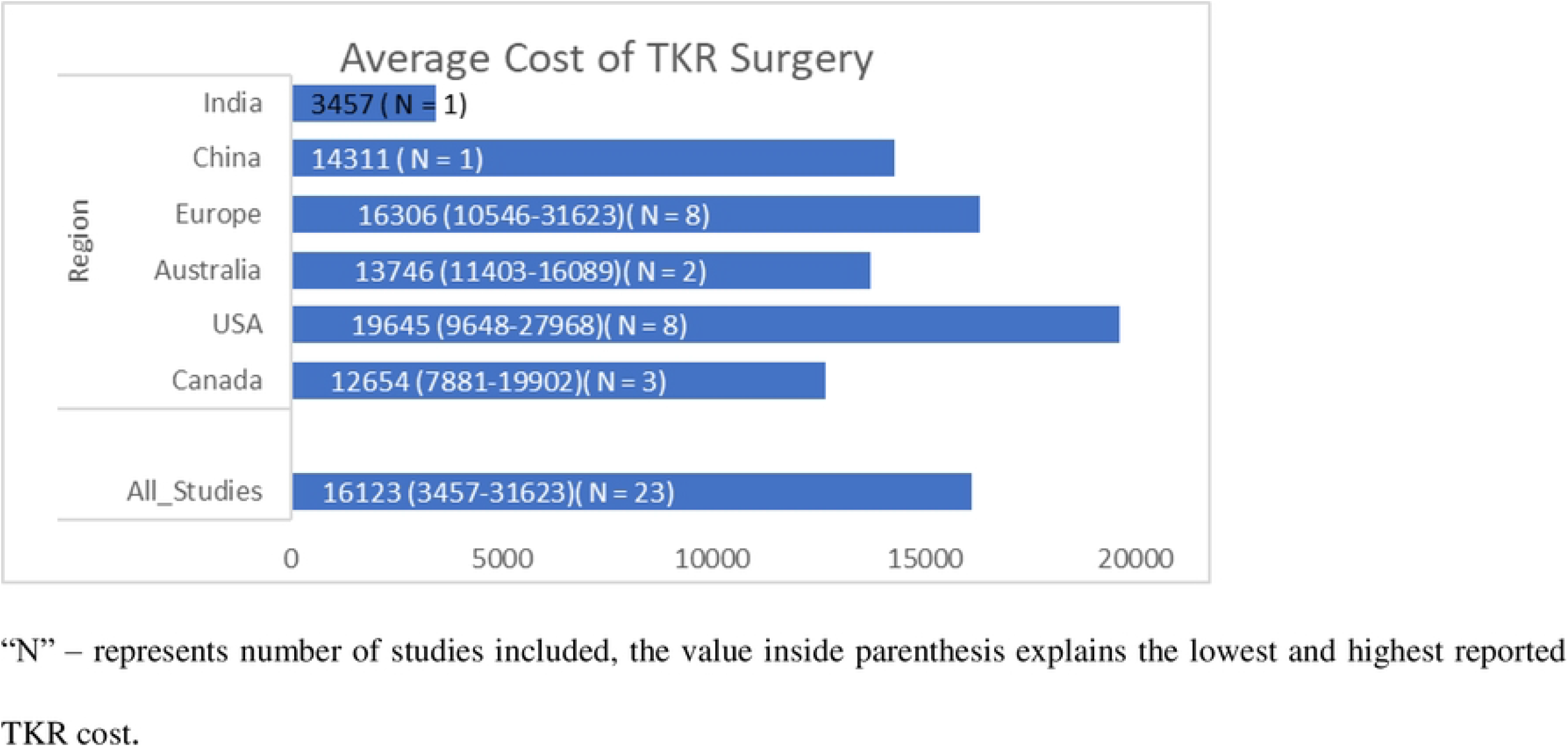
Average cost of TKR surgery.

## Quality of Reporting

CHEERs checklist was used to evaluate the reporting quality of the studies. Nineteen studies were ranked as high, seven as moderate and other three were categorized as low. The proportion of studies that meet the requirements for each of the items has been compiled in S3 File. Only four studies directly stated the comparator name in the title, whereas majority of the studies went into great depth to describe their abstracts. Most of the studies did not report information about setting, analytical model and assumptions, and discount rates. Additionally, neither the findings nor the results of the uncertainty analysis were provided in the papers that were included and only four studies reported conflict of interest.

## Discussion

The main objective of this study was to summarize the evidence on the core modelling specifications on the cost-effectiveness of TKR compared to non-surgical management. Another aim of this study was to synthesize evidence of TKR cost and to compare the variations across different countries through purchasing power parity (PPP) by converting the cost in each study to international dollar ($).

This systematic review assessed the models used in cost utility or cost-effectiveness of the 29 included studies, out of which, sixteen studies reported methodology to determine cost-effectiveness. Eight studies used Markov model (5,7,18,23,24,27,29,37) whereas five studies reported regression model (6,33,34,36,38). One study each reported Marginal structure model(28), discrete simulation model (32) and decision tree analysis (8) to assess the cost-effectiveness of TKR. The regression model explained the variations in cost and outcome of TKR across different age groups and comorbidities. The marginal structure model explained the causal relation between one intervention and health outcome over time. The discrete simulation model studied the cost and outcome of one intervention over the lifetime and found the cost-effectiveness of the intervention. The tree diagram compared the two interventions but with time constraint. On the other hand, the Markov model compared the cost-effectiveness of different interventions over life time and found the incremental cost-effectiveness of one intervention in comparison with other interventions. The findings of the review suggested that the Markov model was most widely used in the analysis of the cost effectiveness of TKR, which is similar to the results of previous economic studies (9,29,37) which showed that Markov model allows a better comparative understanding of cost and health outcome across intervention.

The Markov model uses the outcome (QALY) and cost of two different interventions to calculate ICER. This ICER explains the cost per unit change in effectiveness of new intervention compared to the old intervention. So, the ICER is very easy to understand and is also useful in the decision of policy. The Markov model can use data from real world as well as from assumed data set to calculate ICER value. So, the real-world data can also be used to analyse the budgetary impact of the intervention. Hence, with the help of Markov model, we can calculate the country level effect and cost of any new intervention. The Markov model can also be used in different time frames and study settings. But the path of Markov model is non reversal, so it forgets the last health state of the person.

Of the 29 included studies in our systematic review, 23 studies used direct cost of TKR in the cost effectiveness analysis through Markov model. Of the 23 studies included in the costing analysis, most of them were from US and Europe, only two studies were from the Asian continent; one study from India and another one from China. The direct cost of TKR by each study was reported in their respective country currency. Hence, we converted the cost of TKR reported in all included studies to international dollar ($) through PPP method in order to compare across different countries.

The cost of TKR surgery widely varied across different countries. Another important finding of our review is that the cost of TKR surgery was three to five times higher in developed countries such as USA and UK than developing countries such as India after adjusting cost in PPP terns. These variations in cost were mostly due to the difference in the cost of human resources. Despite the high cost of TKR in developed countries, the TKR surgery was found to be cost effective than non-surgical management for OA knee (23). Similarly, the TKR surgery in developing country like India was also found to be cost effective (7). So, when the economic condition in developing country improves, the cost of TKR surgery may increase but it will still be a cost-effective strategy to treat OA knee.

## Strengths

Our review is first study to suggest a suitable model to estimate the cost effectiveness especially for TKR compared to non-surgical management. Based on our study findings, new studies can utilize Markov model for their analysis and accordingly focus on the appropriate data requirements. Another strength of this review was that our study was the first study to use PPP to convert the cost of TKR surgery into a single currency to compare the cost of TKR across different countries.

## Limitations

Since studies included in our review were from various countries, the generalisability of our findings may be reduced due to the variations in costs and clinical practices across multiple continents. Most of the included studies were from high income countries with the exception of two studies from the low-income countries. Therefore, application research findings to the low-income countries might be a problem. However, the PPP method used to convert currency and appropriate methods as identified by this review may provide insights for uptake of these findings in other country settings.

## Conclusion

The review provides critical insights for guiding model specification for conducting cost effectiveness analysis in TKR as one of the clinical interventions compared to non-surgical management. It is also found that the overall quality of reporting in the cost effectiveness studies is increasing globally however, there are limited number of studies in the low-income countries. This suggests for undertaking more CEA studies in these countries as the burden of OA knee is increasing. This review concluded that Markov model is the most suitable decision model for economic evaluation of TKR and non-surgical management. Across countries, the cost of TKR surgery in a single currency (international $) was found to be lowest in India and highest in a developed country like USA. More studies with high methodological standards in developing countries are recommended.

## Data Availability

All relevant data are within the manuscript and its Supporting Information files.

